# Genetically proxied inhibition of angiotensinogen synthesis is associated with lower cardiovascular risk

**DOI:** 10.64898/2026.03.08.26347887

**Authors:** Panagiotis Zangas, Murad Omarov, Lanyue Zhang, Marios K. Georgakis

## Abstract

**Background and Aims:** Angiotensinogen synthesis inhibitors have shown promising blood pressure-lowering effects in early-stage trials, but their impact on cardiovascular outcomes remains unknown. We investigated associations between genetic variants mimicking angiotensinogen synthesis inhibition and cardiovascular phenotypes.

**Methods:** We developed a genetic proxy for hepatic angiotensinogen synthesis downregulation comprising *AGT* variants that lower liver *AGT* expression (N=1,183) and circulating angiotensinogen levels (N=47,745), selected to mimic the effects of RNA-based angiotensinogen-targeting therapies. Using drug-target Mendelian randomization, we assessed effects on coronary artery disease (210,842 cases, 1,167,328 controls), stroke (110,182 cases, 1,503,898 controls) and heart failure (207,306 cases, 2,151,210 controls), along with vascular endophenotypes and safety outcomes.

**Results:** Mirroring pharmacological angiotensinogen synthesis inhibitors in trials, the *AGT* genetic instrument was associated with lower systolic (SBP, −0.60 [-0.71;-0.48] mmHg) and diastolic blood pressure (DBP, −0.40 [-0.46;-0.33] mmHg), and higher renin and potassium levels. Genetically proxied angiotensinogen synthesis inhibition was associated with lower odds of coronary artery disease (OR per mmHg SBP reduction: 0.954 [0.937-0.972]), stroke (OR: 0.949 [0.928-0.970]) and heart failure (OR: 0.972 [0.957-0.987]) with effect sizes proportional to the SBP-lowering effects of genetic proxies for other renin-angiotensin-aldosterone system drug classes. We found additional associations with lower burden of atherosclerosis, cerebral small vessel disease, and adverse cardiac remodeling on imaging endophenotypes. Aside from hyperkalemia, we detected no links to major safety concerns, including impaired kidney function.

**Conclusions:** Genetic downregulation of angiotensinogen synthesis is associated with lower cardiovascular disease burden without concerning safety signals, supporting the potential of angiotensinogen inhibitors to reduce cardiovascular risk.

**Structured graphical abstract:** *Key Question:* Is there human genetic evidence suggesting that inhibition of hepatic angiotensinogen synthesis can reduce long-term cardiovascular risk?

*Key Finding:* Genetically proxied angiotensinogen synthesis inhibition is associated with lower risk of coronary artery disease, stroke and heart failure, as well as favorable effects on cardiac and cerebrovascular pathologies, without raising major safety concerns. Effect estimates were comparable in magnitude to those observed for genetic proxies of approved RAAS-blocking therapies.

*Take-home Message:* Human genetic evidence supports the hypothesis that angiotensinogen synthesis inhibition may reduce both cardiovascular event risk and chronic subclinical vascular disease burden, providing a strong rationale for prioritizing angiotensinogen inhibitors in cardio- and cerebrovascular outcome trials. Genetically proxied angiotensinogen synthesis inhibition and cardiovascular risk reduction. Graphical overview of the study design. Created in https://www.biorender.com/
AGT: angiotensinogen, pQTL: protein quantitative trait loci, eQTL: expressive quantitative trait loci, LD: linkage disequilibrium, MR: Mendelian Randomization, SBP: systolic blood pressure, DBP: diastolic blood pressure, UKBB: UK Biobank, CAD: coronary artery disease, RAAS: renin-angiotensin-aldosterone system, CVD: cardiovascular disease, ICH: intracerebral hemorrhage, SAH: subarachnoid hemorrhage, cIMT: carotid intima-media thickness, cSVD: cerebral small vessel disease, HF: heart failure, HFrEF: heart failure with reduced ejection fraction, HFpEF: heart failure with preserved ejection fraction, MRI: magnetic resonance imaging, eGFR: estimated glomerular filtration rate, UACR: urinary albumin-to-creatinine ratio, MVP: Million Veteran Program

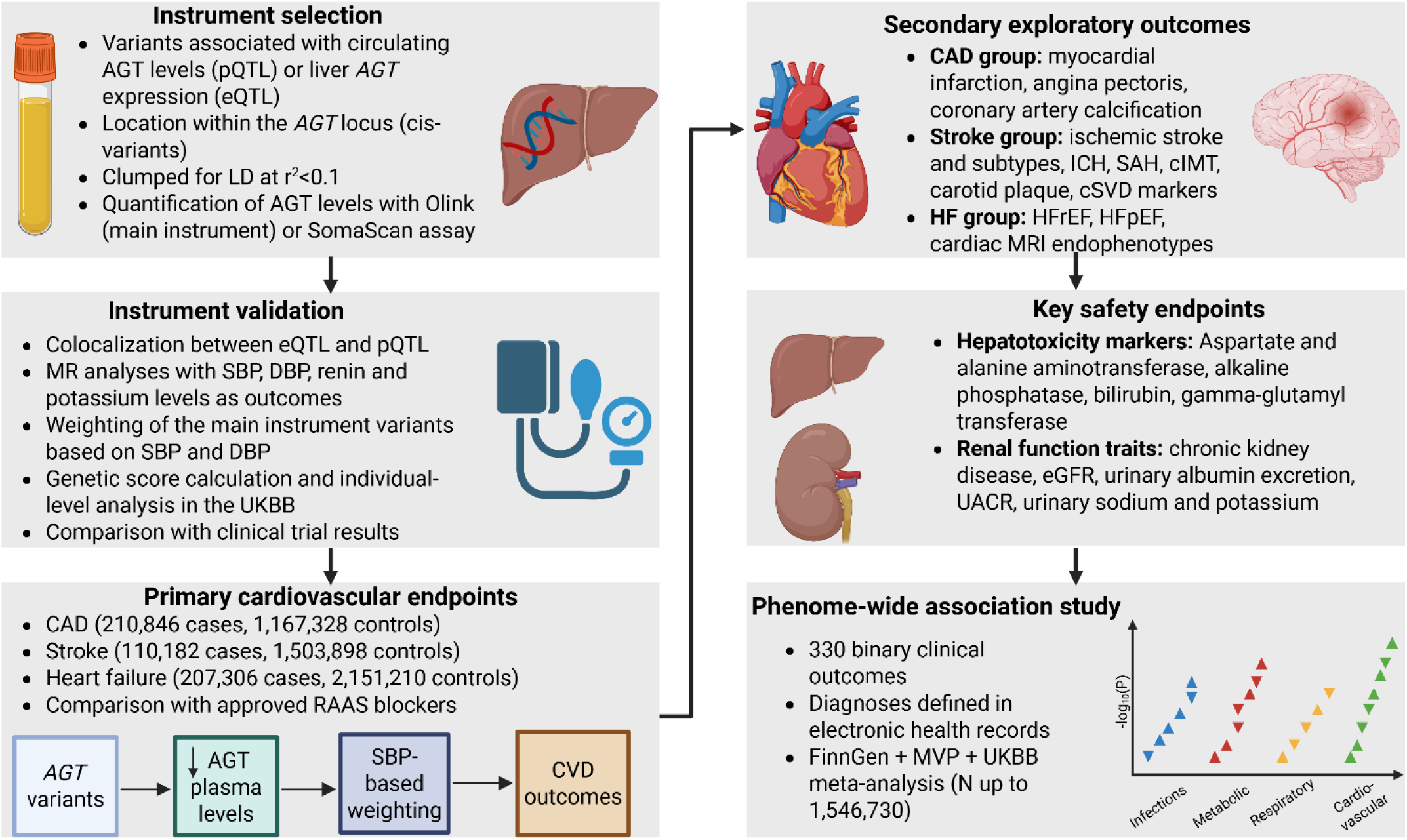

## Introduction

Hypertension is an established risk factor for coronary artery disease (CAD), stroke and heart failure (HF)^1–3^. It is the leading cause of cardiovascular death worldwide^4^ and the primary modifiable risk factor contributing to disability, with high systolic blood pressure (SBP) responsible for approximately 8.4% of total disability-adjusted life years (DALYs)^5^. Despite the wide availability and proven efficacy of different classes of antihypertensive medications, less than half of the adults with hypertension are diagnosed and treated, while only 23% have their blood pressure under control^6^, due in part to suboptimal patient adherence to prescribed daily oral medications^7^. Untreated and poorly treated patients with hypertension are at a higher risk of all-cause and cardiovascular mortality compared to normotensive adults^8^.

The renin-angiotensin-aldosterone system (RAAS) is a key regulator of blood volume, blood pressure and electrolyte balance^9^ and its chronic hyperactivity contributes to the development of atherosclerosis and cardiovascular events^10^. The central role of RAAS in blood pressure regulation has resulted in the development and approval of multiple antihypertensive drug classes targeting its components, namely angiotensin-converting enzyme inhibitors (ACEi)^11^, angiotensin receptor blockers (ARBs)^12^, mineralocorticoid receptor antagonists (MRAs)^13^ and renin inhibitors^14^. Nevertheless, no treatment options targeting angiotensinogen (AGT), the progenitor of the entire angiotensin-peptide family, are currently approved for clinical usage.

To date, three RNA-based therapeutics targeting AGT synthesis have been studied in randomized controlled trials (RCTs), all employing N-acetylgalactosamine (GalNAc) conjugation for liver-specific delivery through the asialoglycoprotein receptor^15^. The antisense oligonucleotide (ASO) IONIS-AGT-L_Rx_ was investigated in phase II RCTs in patients with hypertension^16^ and heart failure (ASTRAAS-HF, NCT04836182), followed by a more potent ASO, ION904,^16^ in patients with uncontrolled hypertension (NCT05314439). Zilebesiran (or ALN-AGT01), a small interfering RNA (siRNA) therapeutic^17^, has been studied in Phase II RCTs as monotherapy for mild- to moderate hypertension (KARDIA-1, NCT04936035)^18^ and as add-on treatment in uncontrolled hypertension (KARDIA-2, NCT05103332)^19^, showing reductions in blood pressure and serum AGT levels. The ongoing KARDIA-3 trial (NCT06272487) is investigating zilebesiran in patients at high cardiovascular risk and inadequately controlled hypertension^20^. Despite promising blood pressure-lowering effects, it remains unknown if AGT synthesis inhibition could translate into cardiovascular risk reduction. ZENITH is the first phase III trial to test the effect of an AGT synthesis inhibitor on cardiovascular outcomes (NCT07181109), but its results are not expected before 2030.

Analyses leveraging human genetic variation in promising drug targets can provide early evidence of potential efficacy and safety for investigational therapeutics^21–23^. Indeed, genetically supported drug targets are 2-3 times more likely to lead to approved drugs^24^. In anticipation of clinical trial results, such data can inform investment and drug development decisions. Here, leveraging this approach, we identified genetic variants in the *AGT* locus mimicking downregulation of hepatic AGT synthesis and used them as instruments in drug-target Mendelian Randomization (MR) analyses to evaluate effects on cardio- and cerebrovascular phenotypes and safety endpoints. We further conducted a phenome-wide association study (PheWAS) to explore potential repurposing opportunities or unexpected adverse effects of AGT synthesis inhibition. Our aim was to assess whether there is genetic support for the hypothesis that hepatic AGT synthesis inhibition could be a promising approach for lowering cardiovascular event rates.

## Methods

This report is compliant with the Strengthening the Reporting of Observational Studies in Epidemiology using Mendelian Randomization (STROBE-MR) statement^25^ **(Supplementary Table 1)**.

### Instrument selection

To proxy the action of AGT synthesis inhibitors, we selected three sets of genetic instruments consisting of genetic variants mimicking their effects on reducing the translation of hepatic *AGT* mRNA and blood AGT levels, as circulating AGT levels are predominantly regulated by hepatic synthesis^26^, and the two traits are known to be positively correlated^27^. Valid instrumental variables must be robustly associated with the exposure (relevance assumption), remain independent of confounders (independence assumption) and influence the outcome only through the exposure (exclusion restriction assumption)^28^.

For our first instrument, we used summary statistics from a UK Biobank (UKBB) GWAS for AGT blood plasma levels (N=47,745, 100% European ancestry)^29^ **(Supplementary Table 2)**. AGT levels in the UKBB were measured with the Olink platform. The variant selection criteria were as follows:

- Location within the *AGT* locus, defined as a window including the *AGT* gene (Chromosome 1, gene base pair location: 230,838,269-230,850,043, based on GRCh37-hg19 genome build) as well as 250kb upstream or downstream to it
- Association with plasma AGT levels at a genome-wide significance level (p<5*10^-8^)
- Lack of strong correlation due to between-variant linkage disequilibrium (clumped at r^2^ <0.1, clumping window = 10,000kb).

To secure stability against the proteomics platform used to quantify AGT levels, as a sensitivity analysis approach, we constructed a second instrument using a GWAS of AGT plasma levels quantified with the SomaScan multiplex aptamer assay among 35,559 Icelandic individuals (deCODE genetics)^30^ **(Supplementary Table 2)**. The variant selection criteria for this instrument were the same as above.

Finally, to directly proxy mRNA synthesis inhibition in liver, we derived *AGT* hepatic expressive quantitative trait loci (eQTL) from a liver eQTL meta-analysis of 4 studies (N=1,183, 100% European ancestry)^31^ **(Supplementary Table 2)**. We selected the variants based on the following criteria:

- Cis-eQTL, located within 1Mb of the *AGT* gene transcription start site
- Association *with AGT* expression levels at a p-value < 1*10^-5^, equivalent to significant false discovery rate (FDR)- adjusted p-values in the original publication^31^
- Clumping at r^2^ <0.1, (clumping window = 10,000kb).

The hepatic eQTL instrument was also utilized in sensitivity analyses, given its limited strength compared to the UKBB circulating AGT instrument. For all the instruments and each variant separately, we calculated F-statistics and the proportion of variance they explained in plasma AGT levels or hepatic *AGT* gene expression^32^. **(Supplementary Table 3)**.

### Instrument validation and weighting

To validate the relationship between liver *AGT* expression and circulating AGT levels and to identify potential common causal variants, we performed a colocalization analysis in the *AGT* genomic region between the variants that modify hepatic gene expression and blood protein levels.

Angiotensin II regulates renin secretion through a negative feedback loop^33^, leading to an increase in renin levels in response to a decrease in angiotensin II levels caused by AGT inhibition. Both *Agt* gene modification and protein synthesis inhibition using siRNA largely increased renin levels in mice^34,35^. Moreover, individuals treated with IONIS-AGT-L_Rx_ demonstrated higher renin levels than the placebo group in phase II hypertension trials, although statistical power to determine significant differences was not sufficient^16^. Based on this evidence, we performed a MR analysis with circulating renin levels (N=21,758, 100% European ancestry)^36^ **(Supplementary Table 2)** as a positive control. We additionally assessed the instrument effects on blood potassium levels (N=603,756, 70% European ancestry)^37^ **(Supplementary Table 2)**, since hyperkalemia is a common side effect of drugs inhibiting RAAS components^38^ and mild, transient hyperkalemia was observed in a greater proportion of patients receiving zilebesiran than placebo in the KARDIA-1 and KARDIA-2 trials^18,19^.

To determine whether the genetic instruments have blood pressure-lowering effects, we explored their effects on systolic blood pressure (SBP) and diastolic blood pressure (DBP). Data for SBP and DBP were derived from a GWAS meta-analysis of the UKBB and the International Consortium for Blood Pressure (ICBP) (N=757,601, 100% European ancestry)^39^ **(Supplementary Table 2)**. For the UKBB AGT instrument, we performed additional SBP and DBP analyses excluding UKBB population, using GWAS summary statistics from the Million Veteran Program (MVP, N=609,479 for SBP, N=609,354 for DBP, 70% European ancestry)^37^ **(Supplementary Table 2)**.

Moreover, we used GWAS summary statistics for left-handedness (N=1,766,671, 100% European ancestry)^40^, a neurodevelopmental trait with no known connection to blood pressure or the RAAS, as a negative control **(Supplementary Table 2)**.

We weighted the variants present in both the UKBB AGT-lowering instrument and the blood pressure GWAS according to their effect on SBP and DBP, ensuring that the MR estimates reflect interpretable BP-lowering effects in accordance with pharmacological AGT inhibition rather than the general genetic perturbation in the *AGT* locus.

### Individual-level analysis in the UK Biobank and comparison with clinical trial results

To assess the effects of our instruments on circulating AGT levels, SBP and DBP, we analyzed individual-level data from the UKBB, a UK-wide prospective cohort of 503,317 adults aged 39-73, recruited between 2006 and 2010^41^. We performed the analysis on 384,908 individuals with available genetic data, blood pressure values and no history of antihypertensive medication use and on 44,428 individuals with available genetic data and AGT levels in plasma. Using the clumping and thresholding method^42^, we calculated genetic scores for circulating AGT levels, SBP and DBP, and calculated differences between the top and bottom percentiles of each distribution. For the computation of the blood pressure-related scores, the variants were weighted based on their effects on SBP and DBP. The scores were calculated using *plink 2.00a*^43^.

Consequently, we examined proportionality between genetically proxied and pharmacological (300mg zilebesiran every six months, dose currently being administered in the ZENITH trial) AGT synthesis inhibition by comparing the KARDIA-1 results^18^ with the differences in mean SBP and DBP between the top and bottom 1% of the respective genetic score distribution in the UKBB, per 1-standard deviation (SD) unit decrease in circulating AGT levels.

### Cardio- and cerebrovascular outcomes

Our primary outcomes included the clinical endpoints of CAD, stroke and HF, traits being endpoints in the ZENITH trial and strongly connected with elevated blood pressure^3,44^. We obtained the effects of the variants comprising our genetic instruments on these outcomes from publicly available GWAS summary statistics. For CAD we used data from a meta-analysis of the CARDIoGRAMplusC4D consortium, UKBB and Biobank Japan (210,842 cases, 1,167,328 controls, ∼85% European ancestry)^45^, for stroke from the GIGASTROKE study (110,182 cases, 1,503,898 controls, 81% European ancestry)^46^ and for HF from a GWAS meta-analysis of eight separate cohorts/consortia (207,306 cases, 2,151,210 controls, 80% European ancestry)^47^ **(Supplementary Table 2)**. We also assessed the robustness of our main instrument in replication analyses using data from GWAS meta-analyses excluding UKBB participants, namely CARDIoGRAMplusC4D for CAD (60,801 cases, 123,504 controls)^48^, MEGASTROKE for stroke(67,162 cases and 454,450 controls)^49^ and the HERMES consortium for HF (40,805 cases, 542,362 controls)^50^ **(Supplementary Table 2)**.

As secondary exploratory outcomes we explored associations with a wide range of cardio- and cerebrovascular clinical and imaging outcomes. We classified these outcomes in three groups (CAD, stroke and HF group). In the first category (CAD group) we used GWAS summary statistics for myocardial infarction (MI, 61,505 cases, 577,716 controls)^51^, angina pectoris (44,032 cases, 586,604 controls)^52^ and coronary artery calcification (CAC, N=35,776)^53^ **(Supplementary Table 2)**. We divided the stroke group into two subcategories. The first subgroup included stroke subtypes from the GIGASTROKE study^46^, namely ischemic stroke (86,668 cases, 1,503,898 controls), large artery stroke (9,219 cases, 1,503,898 controls), small-vessel stroke (13,620 cases, 1,503,898 controls) and cardioembolic stroke (12,790 cases, 1,503,898 controls), as well as intracerebral hemorrhage (ICH, 3,391 cases, 623,600 controls)^52^, subarachnoid hemorrhage (SAH, 2,896 cases, 623,584 controls)^52^ and vascular dementia (3,892 cases, 466,606 controls) ^54^**(Supplementary Table 2)**.The second subgroup consisted of cerebrovascular imaging markers related to carotid atherosclerosis and cerebral small vessel disease (cSVD), specifically carotid intima-media thickness (cIMT, N=71,128)^55^, ultrasound-defined carotid plaque presence (21,540 cases, 26,894 controls)^55^, white matter hyperintensity (WMH) volume (N=48,454)^56^, mean diffusivity (N=34,334) and fractional anisotropy (N=35,004)^57^, basal ganglia enlarged perivascular spaces (9,189 cases, 30,811 controls)^58^, and cerebral microbleeds (3,556 cases, 22,306 controls)^59^ **(Supplementary Table 2)**. The HF group was comprised of the two main HF subtypes, namely HF with reduced ejection fraction (HFrEF, 43,187 cases, 579,600 controls) and HF with preserved ejection fraction (HFpEF, 26,634 cases, 589,369 controls)^37^, and N-terminal pro-B natriuretic peptide (NT-proBNP, N=21,758)^36^. Furthermore, we studied associations with cardiac magnetic resonance imaging (MRI)-derived HF endophenotypes: left ventricular end-diastolic volume (LVEDV), end-systolic volume (LVESV), mass (LVM) and mean wall thickness (LVmeanWT, N up to 36,083, 100% European or unknown ancestry)^60^ **(Supplementary Table 2)**. LVESV, LVEDV and LVM were indexed to body surface area.

### Safety-related outcomes

To assess potential safety concerns of angiotensinogen inhibitors, we additionally explored renal and hepatic outcomes, as they were the main safety endpoints of interest in clinical trials ^17–19^. The outcomes we examined were alanine aminotransferase (ALT, N=494,681), aspartate aminotransferase (AST, N=493,058), bilirubin (N=467,170), gamma-glutamyl transferase (gamma-GT, N=477,575) and alkaline phosphatase (ALP, 463,178) levels in blood serum^52^, as well as estimated glomerular filtration rate (eGFR, N=1,201,909)^61^, chronic kidney disease (64,164 cases, 561,055 controls)^62^, urinary albumin to creatinine ratio (UACR, N=564,257)^63^, urinary albumin excretion (N=382,500)^64^, urinary sodium (N=446,237)^65^ and urinary potassium (N=446,230)^65^. Data sources for the safety-related outcomes can be found in **Supplementary Table 2.**

### Comparison with approved RAAS blockers

We investigated the efficacy of genetically proxied AGT synthesis inhibition on the three primary outcomes compared to genetic proxies of already approved RAAS inhibitors per BP-lowering unit. To achieve this, we constructed an instrument for every target (*AGT* for angiotensinogen inhibitors, *ACE* for ACEi, *AGTR1* for ARBs, *NR3C2* for MRAs and *REN* for renin inhibitors), based on the direct effects of the variants on SBP. We then performed two-sample MR analyses and assessed presence of heterogeneity between the effect estimates using the Cochran’s Q test. The instrument selection criteria for this analysis were as follows:

- Location within a window including the target gene and 250kb up- and downstream to it
- Association with SBP at a p<10^-5^, as genome-wide significant variants were not available for most targets beyond *AGT*. In the *NR3C2* locus no variants were found despite the less stringent threshold, resulting in the exclusion of this target from the analysis
- Clumping at r^2^<0.1, clumping window = 10,000kb

### Statistical analysis

#### Mendelian Randomization

We performed two-sample MR analyses using the *TwoSampleMR* R package (version 0.6.19). We used fixed-effects Inverse variance weighted (IVW) MR as our primary analysis method ^66^. The Cochran Q test was used to assess presence of heterogeneity (p<0.05 indicates heterogeneity between the variants) ^67^. MR Egger regression and weighted median MR were used as sensitivity analyses, where applicable. The weighted median method provides a consistent estimate of the causal effect, as long as at least 50% of the instrumental variables are valid ^68^. MR Egger regression assumes that the effect of each instrument with the exposure is independent of any pleiotropic effect^69^. An Egger intercept significantly different from zero (p<0.05) provides an indication of horizontal pleiotropy. The Wald ratio method was applied if the instrument consisted of only one variant^70^. The main analyses for primary and secondary cardio- and cerebrovascular outcomes, were weighed based on the SBP-lowering effects of the AGT level instrument (1 mmHg-reduction) to derived interpretable MR estimates comparable to those expected from clinical trials. In sensitivity and safety outcome analyses, we scaled the instrument based on its effects on plasma AGT levels (1-standard deviation (SD) reduction). In further sensitivity analyses, we used the SomaScan-based instrument for circulating AGT levels and the instrument proxying hepatic *AGT* mRNA expression (also 1 SD-reduction). The MR estimates (beta coefficients) for the cardiac MRI endophenotypes were scaled to represent 1-SD unit change in each measurement. The statistical significance threshold was set at p<0.0167 adjusting for multiple comparisons for our three main outcomes using the Bonferroni approach, while 0.0167<p<0.05 indicated nominal significance. For the secondary exploratory and safety-related outcomes the threshold was set at p<0.05. All p-values are two-sided.

#### Colocalization analysis

To ensure that the signal captured for circulating AGT levels was driven by effects on hepatic *AGT* expression, we performed a colocalization analysis in a Bayesian framework^71^, using the *coloc* R package. We tested the posterior probability of several hypotheses, in particular H0 (no association for either trait), H1 (association only with AGT levels, UKBB summary statistics), H2 (association only with gene expression), H3 (association with both traits through two distinct variants) and H4 (association with both traits through the same variant)^71^. We set the prior probabilities at 10^-4^ for a variant associated with AGT levels, 10^-4^ for a variant associated with gene expression, and 10^-5^ for a variant associated with both traits. A posterior probability of H4 >80% provides strong evidence for colocalization^71^. We used common (minor allele frequency >1%) variants for each trait within the region that defined the cis-eQTL window in the original publication^31^, namely within +/- 1Mb of the gene transcription start site.

### Phenome-wide association study

We conducted a phenome-wide MR analysis using summary statistics from a GWAS meta-analysis of FinnGen^72^, MVP^37^ and UKBB^73^. This study comprises 330 clinical outcomes, based on diagnoses extracted from electronic health records, with the sample size ranging from 730,332 to 1,546,730 participants. From the original outcome list, we excluded four traits with less than 1000 cases, leading to a set of 326 binary phenotypes. Similar to the cardiovascular outcome analyses, IVW MR was selected as the primary analytical method. We applied the Benjamini-Hochberg method in the initial results to correct for multiple comparisons, considering an FDR-adjusted p<0.05 as statistically significant. To further validate the findings, we performed weighted median MR and MR Egger regression as sensitivity analyses. The Olink-based instrument was used for this analysis, and the derived odds ratios correspond to 1-SD reduction in plasma AGT levels.

### Ethics

The UK Biobank has obtained approval from the Northwest Multi-Center Research Ethics Committee (REC reference for UK Biobank is 11/NW/0382). All participants have provided written informed consent. The publicly available GWAS summary statistics are provided by studies that received ethical approval and participant consent for both the analysis and distribution of summary-level data, as described in the original publications.

## Results

### Genetic instruments

A total of 46 variants in the *AGT* locus meeting the selection criteria were included in the primary genetic instrument. All F-statistics were >10, indicating sufficient instrument strength^32^. The instrument explained 15.2% of the variance in plasma AGT protein levels **(Supplementary Table 3)**. The liver eQTL and SomaScan-based complementary instruments consisted of 2 and 18 variants, respectively, explaining 3.8% of hepatic *AGT* expression and 4.2% of AGT plasma levels variance. **(Supplementary Table 3)**. There was strong evidence of colocalization between gene and protein expression in the *AGT* locus (PPH4 =99%), suggesting the presence of a genetic common signal in the *AGT* locus for liver RNA expression and circulating levels and also genetically supporting that circulating AGT levels are predominantly regulated by hepatic synthesis.

### The genetic instrument proxies pharmacological angiotensinogen synthesis inhibition

In the UKBB, differences of 0.814 mmHg in mean SBP (95% CI: −0.002; 1.629), 0.703 mmHg in mean DBP (95% CI: 0.252; 1.164) and 0.31 SD units (95% CI: 0.27; 0.34) in mean blood AGT were observed between the top and bottom 1% of the genetic score distributions **(Figure 1, Supplementary Table 4)**. These effects were comparable to zilebesiran effects in the KARDIA-1 trial, after scaling the estimates to represent 1-SD reduction in circulating AGT (Genetic effect: 2.6 mmHg SBP and 2.3 mmHg DBP reduction, Drug effect: 3.3 mmHg SBP and 2 mmHg DBP reduction).

**Figure 1.**
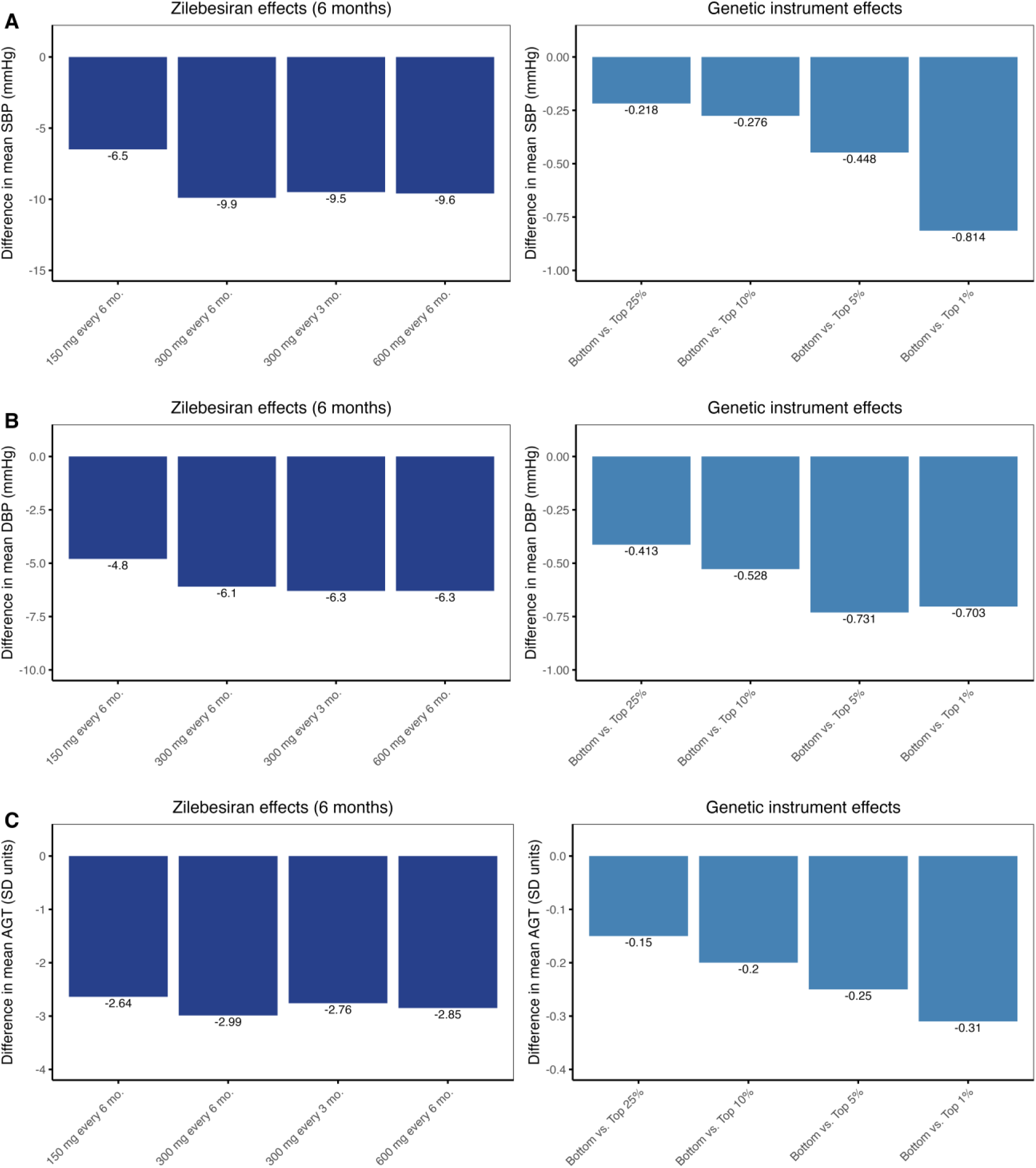
Comparison of drug and genetic effects on blood pressure. Bar plots showing the results of the individual-level data analysis in the UK Biobank, demonstrating the differences in mean SBP **(A)**, DBP **(B)** and circulating AGT **(C)** between the bottom and top 1%, 5%, 10% and 25% of the genetic score distribution derived from the blood pressure- and AGT-lowering instruments and comparisons with the 6-month zilebesiran effects in the KARDIA-1 trial. SBP-systolic blood pressure, DBP – diastolic blood pressure, AGT – angiotensinogen, SD – standard deviation.

Genetically predicted reduction in AGT levels was further associated with increases in renin (standardized beta per 1-SD reduction in AGT levels: 0.089, 95% CI: 0.049; 0.129, p=1.1*10^-5^), and potassium levels (standardized beta: 0.026, 95% CI: 0.015; 0.036, p=7.6*10^-7^), as well as decreases in SBP (beta: −0.597 mmHg, 95% CI: −0.714;-0.479, p = 3*10^-23^) and DBP (beta: −0.396 mmHg, 95% CI: −0.463; −0.329, p=1.1*10^-30^) **(Figure 2, Supplementary Table 5).** The directionality and significance of the effects persisted when using blood pressure data derived from a non-UKBB population, minimizing potential bias from population overlap between the exposure and outcome datasets **(Supplementary Table 5).** The complementary instruments were also associated with lower SBP, lower DBP and higher potassium levels, but not with renin levels **(Supplementary Figures 1 and 2, Supplementary Table 5).** In the negative control analysis, no significant associations were observed between either of the three instruments and left-handedness **(Supplementary Table 5).**

**Figure 2.**
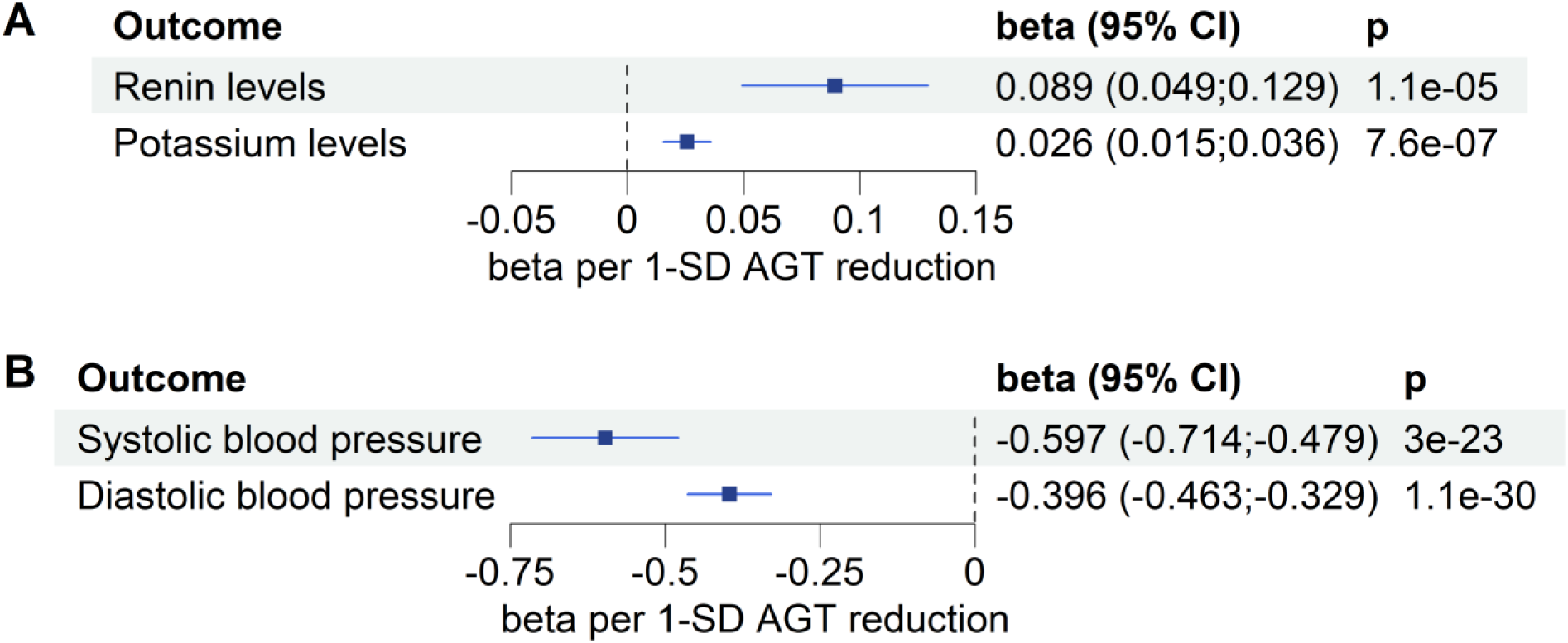
Positive control analysis for the main protein-lowering instrument. Forest plots depicting the IVW MR effects of the Olink plasma AGT instrument on **(A)** circulating renin and potassium levels, **(B)** systolic and diastolic blood pressure. CI: confidence intervals, SD: standard deviation, AGT: angiotensinogen.

### Angiotensinogen genetic perturbation is associated with lower cardiovascular risk

Genetically proxied angiotensinogen synthesis inhibition was significantly associated with CAD (OR per 1mmHg SBP reduction: 0.954, 95% CI: 0.937; 0.972, p=3.8*10^-7^), stroke (OR: 0.949, 95% CI: 0.928; 0.970, p=2.1*10^-6^) and HF (OR: 0.972, 95% CI: 0.957; 0.987, p=2.4*10^-4^) **(Figure 3, Supplementary Table 6)**. There was no evidence of between-variant heterogeneity for CAD and stroke, and despite evidence for heterogeneity for HF (p<0.05), the MR estimates were largely consistent in sensitivity analyses robust to inclusion of pleiotropic variants. In particular, the weighted median estimates were significant for CAD (OR: 0.960, 95% CI: 0.933; 0.989, p=0.006) and stroke (OR: 0.950, 95% CI: 0.921; 0.981, p=0.001), and nominally significant for HF (OR: 0.974, 95% CI: 0.951; 0.997, p=0.028). The effect magnitude was also similar in the MR Egger regression analysis (OR: 0.954, 95% CI: 0.926; 0.983, p=0.004 for CAD, OR: 0.937, 0.904; 0.972, p=0.001 for stroke, OR: 0.971, 95% CI: 0.944; 0.999, p=0.046 for HF), without evidence of significantly different from zero intercepts, which would provide an indication of horizontal pleiotropy **(Figure 3, Supplementary Table 6).** The results further remained consistent when using outcome datasets not including UKBB participants **(Supplementary Table 6),** as well as when pursuing the analyses using the complementary instruments directly proxying (Olink- or SomaScan-based) plasma AGT levels or liver mRNA *AGT* expression **(Supplementary Figure 3, Supplementary Table 6).**

**Figure 3.**
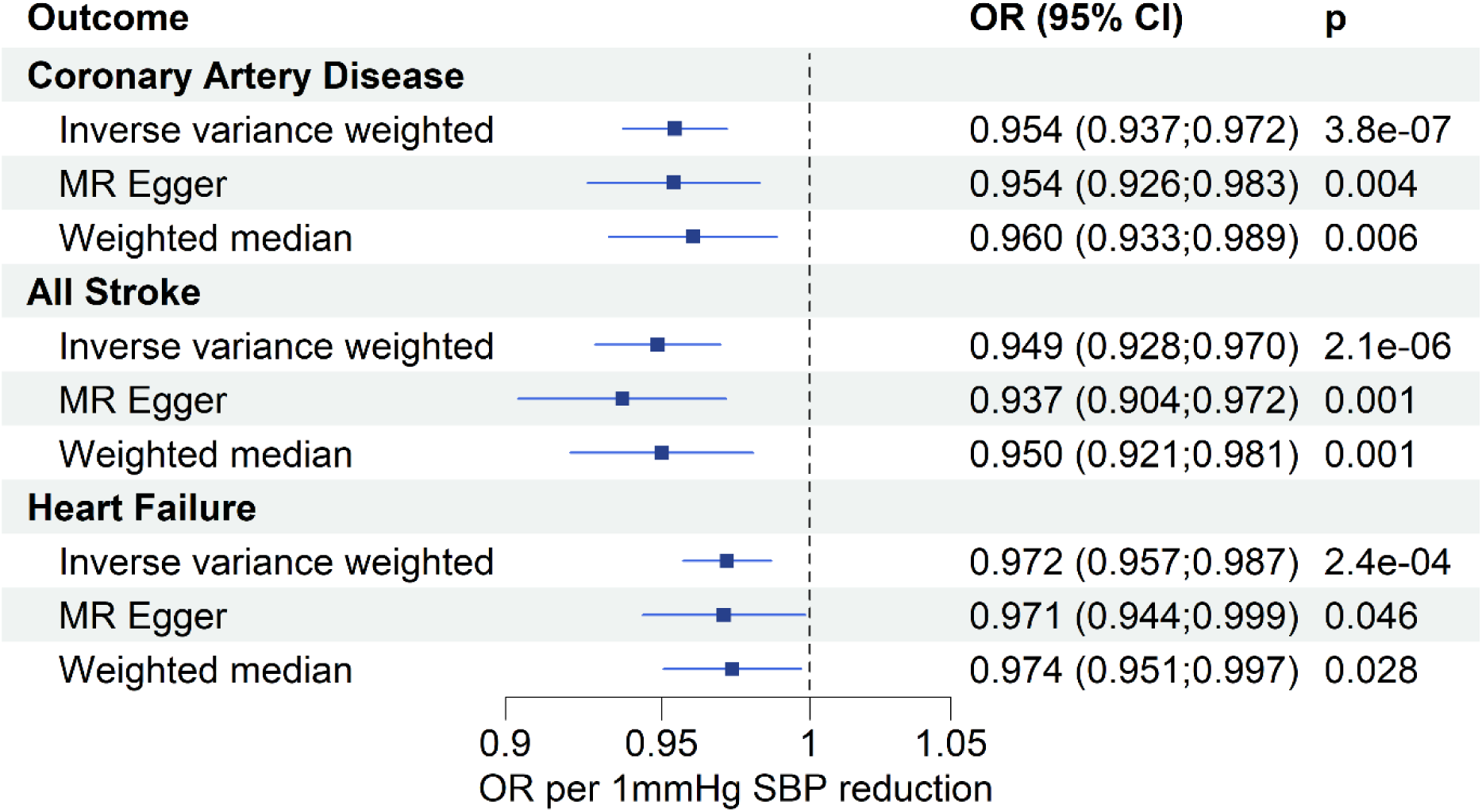
Mendelian Randomization results for primary outcomes. Forest plot showing the IVW, MR-Egger and weighted median MR effects of the SBP-weighted AGT instrument on CAD, stroke and heart failure risk. OR – odds ratio, CI – confidence intervals, SBP – systolic blood pressure.

In a subsequent analysis we compared the SBP-lowering effects of genetically proxied AGT synthesis inhibition on cardiovascular outcomes to those of genetic proxies of other RAAS-blocking therapies (3 variants in *ACE*, 1 variant *in AGTR1*, and 2 variants in *REN*). We found no evidence of heterogeneity across the drug target instruments (all Cochran Q-derived p >0.1) for any of the three main outcomes (CAD, stroke, HF, **Figure 4**). Notably though, the *AGT* SBP-lowering instrument was the only one demonstrating associations with all three cardiovascular outcomes at a p<0.05 **(Figure 4, Supplementary Table 7).**

**Figure 4.**
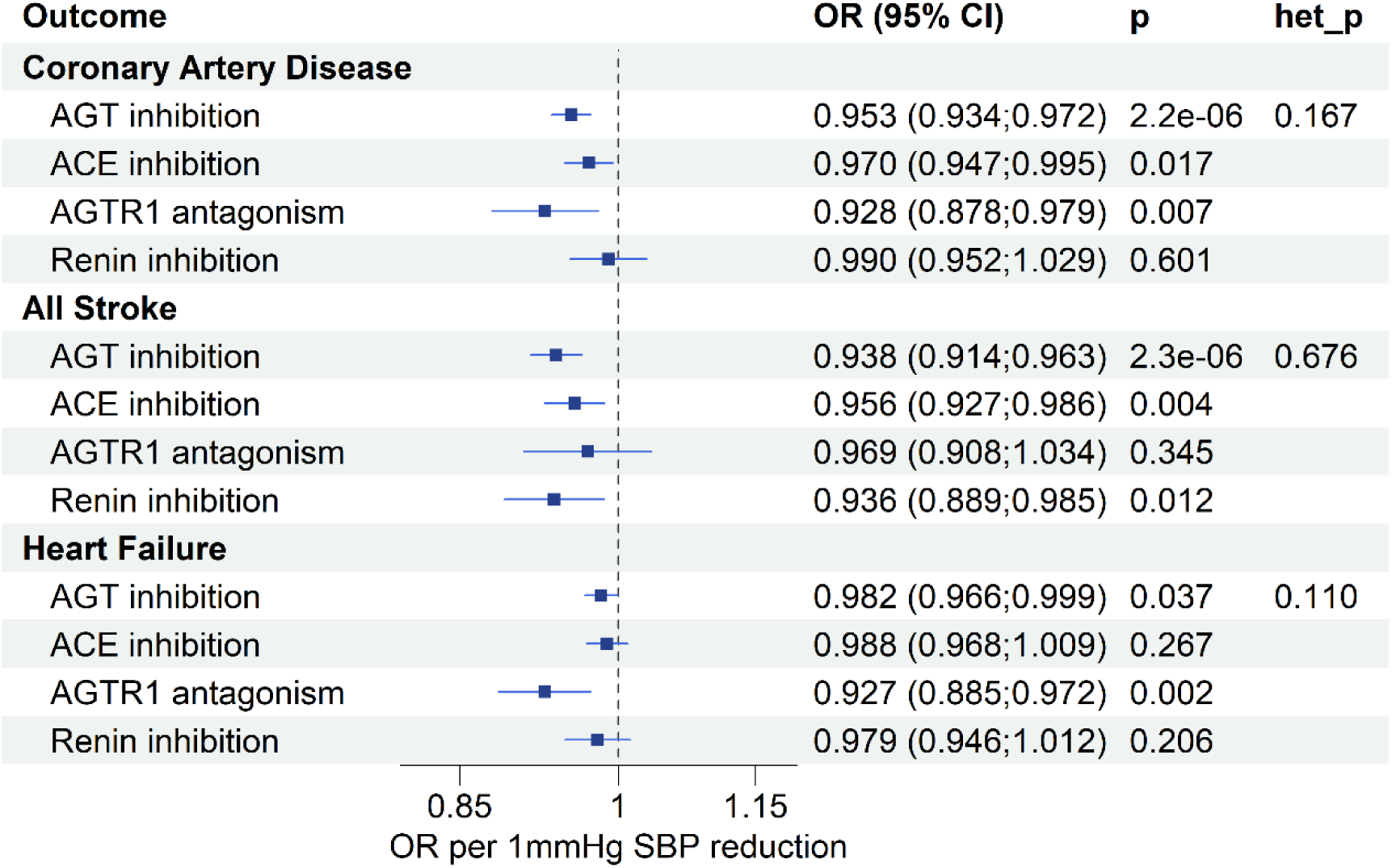
Mendelian Randomization results of instruments proxying RAAS inhibition in different levels. Forest plot depicting the IVW MR effects of the four genetic instruments proxying RAAS-blocking drugs on coronary artery disease, stroke and heart failure. OR: odds ratio, CI: confidence intervals, AGT: angiotensinogen, ACE: angiotensin-converting enzyme, AGTR1: angiotensin II receptor type I, SBP: systolic blood pressure

### Angiotensinogen genetic perturbation is associated with additional markers of atherosclerosis, cerebral small vessel disease, and adverse cardiac remodeling

We next explored associations with secondary clinical endpoints, as well as endophenotypes of atherosclerosis, cerebrovascular pathologies, and adverse cardiac remodeling. With respect to the CAD subgroup of outcomes, we observed associations of genetically proxied AGT synthesis inhibition with MI (OR per 1 mmHg SBP reduction: 0.923, 95% CI: 0.896; 0.951, p=1.2*10^-7^), angina pectoris (OR: 0.937, 95% CI: 0.907; 0.967, p=6.7*10^-5^) and CAC (beta: −0.171, 95% CI: −0.252; −0.089, p=4.1*10^-5^) **(Figure 5, Supplementary Table 8).** Among the clinical cerebrovascular outcomes, our main instrument was correlated with lower odds of ischemic (OR per 1mmHg SBP reduction: 0.946, 95% CI: 0.923; 0.970, p=8.6*10^-6^), cardioembolic (OR: 0.934, 95% CI: 0.877; 0.994, p=0.033), small-vessel stroke (OR: 0.941, 95% CI: 0.887; 0.999, p=0.047) and SAH (OR: 0.883, 95% CI: 0.791; 0.986, p=0.027) **(Figure 5, Supplementary Table 9).** With respect to brain imaging features, the main instrument was associated with reduced mean diffusivity (beta: −0.045, 95% CI: −0.071; −0.019, p=6.2*10^-4^) and lower WMH burden (beta: −0.042, 95% CI: −0.067; −0.017, p=0.001) in magnetic resonance imaging, and lower risk of carotid plaque presence in ultrasound (OR: 0.933, 95% CI: 0.871; 0.999, p=0.049) **(Figure 5, Supplementary Table 9)**. All three complementary instruments were associated with lower risk of MI, ischemic stroke and reduced WMH volume **(Supplementary Figures 4-6, Supplementary Tables 8-9)**. In the HF group, none of the instruments were associated with HFrEF, HFpEF or NT-proBNP **(Figure 5, Supplementary Figures 4-6, Supplementary Table 10)**. With respect to the cardiac MRI endophenotypes, the SBP-weighted instrument was associated with decreases in LVEDV (standardized beta per 1mmHg SBP reduction: −0.041, 95% CI: −0.067; −0.014, p=0.003), LVmeanWT (standardized beta: −0.042, 95% CI: −0.060; −0.023, p=7.6*10^-6^) and LVM (standardized beta: −0.068, 95% CI: −0.090; −0.046, p=1.8*10^-9^) **(Figure 5, Supplementary Table 10)**. The results on secondary endpoints were largely stable in sensitivity analyses using alternative MR methods or the complementary instruments **(Supplementary Figures 4-6, Supplementary Tables 8-10).**

**Figure 5.**
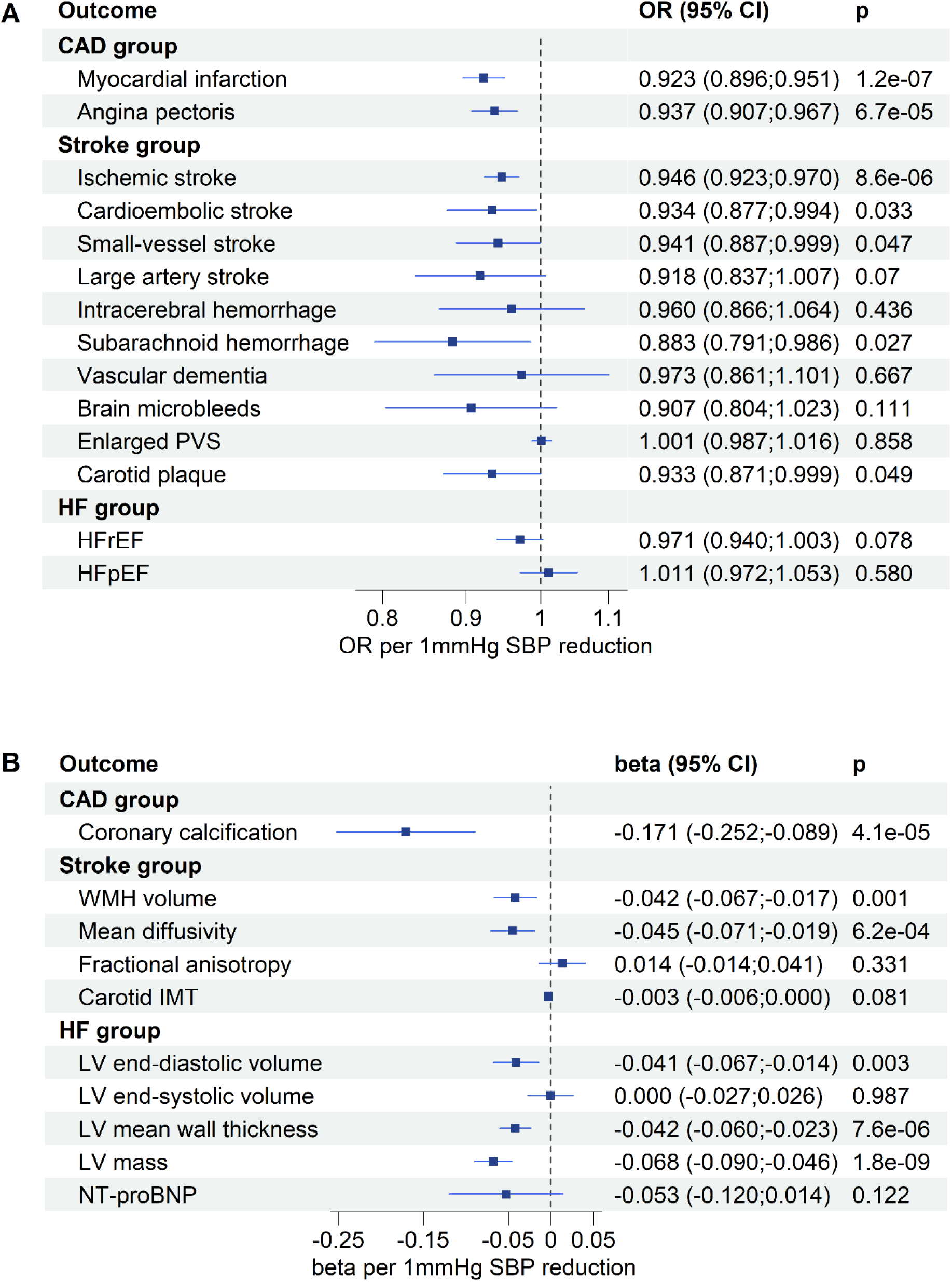
Mendelian Randomization results for secondary outcomes. Forest plots showing the IVW MR effects of the SBP-weighted angiotensinogen instrument on binary **(A)** and continuous **(B)** outcomes of the CAD, stroke and HF subgroups. The beta coefficients of the HF group continuous outcomes were scaled to represent 1-SD changes in each measurement. OR – odds ratio, CI – confidence intervals, SBP – systolic blood pressure, CAD – coronary artery disease, PVS – perivascular spaces, HF – heart failure, HFrEF – heart failure with reduced ejection fraction, HFpEF – heart failure with preserved ejection fraction, WMH – white matter hyperintensities, IMT – intima-media thickness, LV – left ventricle, NT-proBNP – N-terminal pro-B natriuretic peptide.

### Safety-related outcomes

In addition to the above-mentioned increase in blood potassium levels, genetically proxied AGT synthesis inhibition was associated with an increase in urinary sodium (beta per 1-SD reduction in plasma AGT: 0.006, 95% CI: 0.001; 0.010, p=0.018), as expected given the mineralocorticoid role of the RAAS. Among potential safety signals, we could not find any effects on markers of reduced kidney function or kidney injury. On the contrary, there was an association with lower risk of CKD (OR per 1-SD reduction: 0.966, 95%CI: 0.936; 0.998, p=0.035). Exploring on-target liver injury effect, we found an isolated association with higher AST levels (beta per 1-SD reduction: 0.012, 95%CI: 0.004; 0.02, p=0.003), which was not accompanied by any effects on ALT, ALP, gamma-GT or bilirubin levels **(Figure 6, Supplementary Table 11)**. Across the complementary instruments, no significant effects were detected for genetically proxied liver *AGT* expression **(Supplementary Figure 7, Supplementary Table 11),** whereas the SomaScan-based plasma AGT instrument was associated with an increase in gamma-GT levels, and decreases in eGFR, UACR and urinary albumin excretion **(Supplementary Figure 8, Supplementary Table 11).**

**Figure 6.**
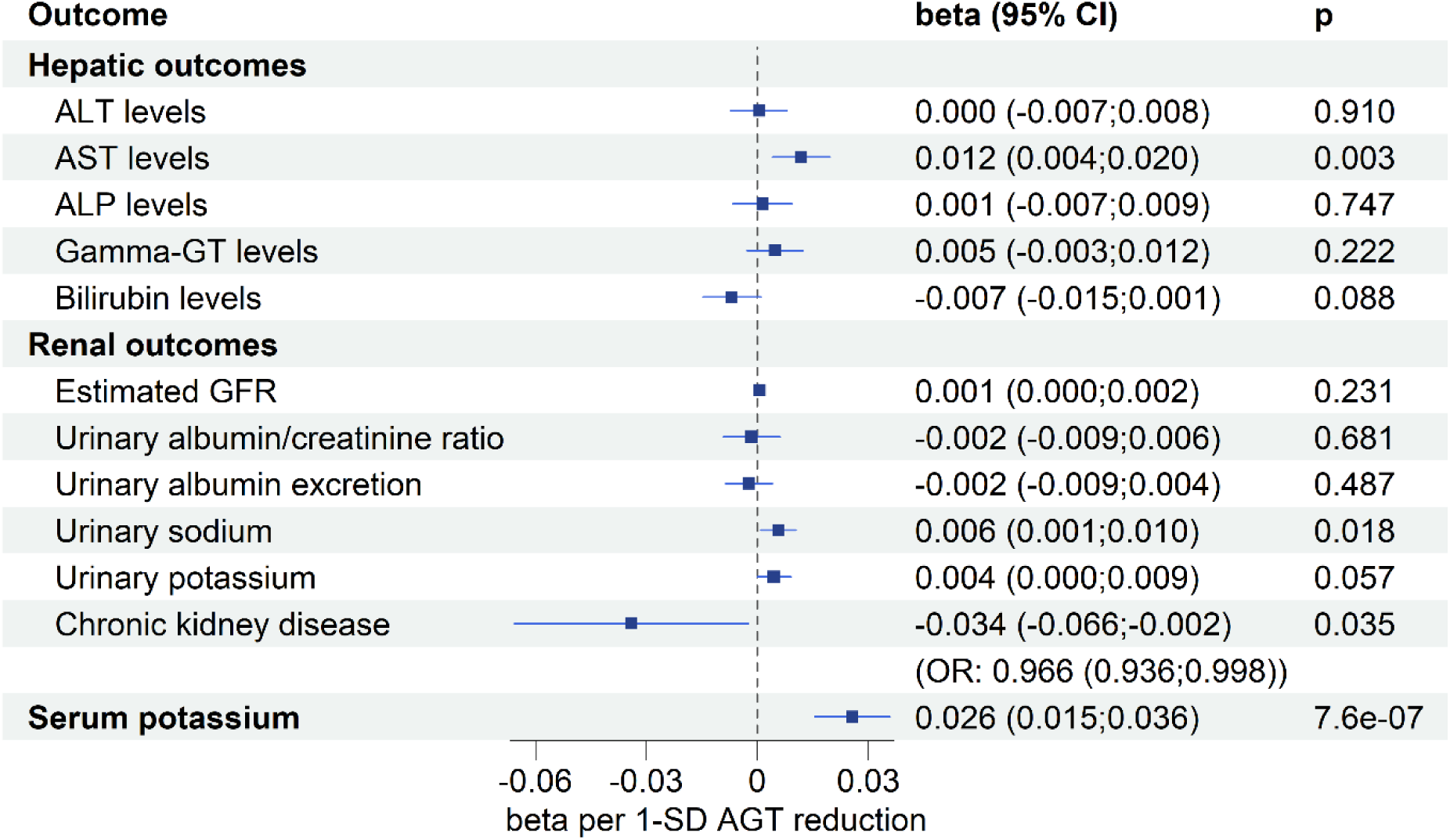
Mendelian Randomization results for safety-related outcomes. Forest plot showing the IVW MR effects of the main angiotensinogen-lowering instrument on liver- and kidney related parameters as well as serum potassium. OR – odds ratio, CI – confidence intervals, GFR – glomerular filtration rate, SD – standard deviation, AGT – angiotensinogen, ALT – alanine aminotransferase, AST – aspartate aminotransferase, ALP – alkaline phosphatase, Gamma-GT - gamma-glutamyl transferase.

### Phenome-wide analyses highlight repurposing opportunities and reveal no major safety signals

To explore associations across the disease spectrum that could inform repurposing strategies or highlight unexpected safety signals, we conducted a PheWAS meta-analysis of 326 clinical outcomes across UKBB, FinnGen, and MVP (N up to 1,546,730). After multiple testing correction (FDR-adjusted p<0.05), genetic downregulation of AGT synthesis was associated with 40 clinical outcomes (**Figure 7, Supplementary Table 12)**, 33 showing reduced odds (potential efficacy signals) and 7 showing increased odds (potential safety signals). 13 of the significant outcomes reflected associations between genetically proxied downregulation of AGT synthesis and lower odds of circulatory system-related endpoints related to the circulatory system, largely validating the cardiovascular results of our analyses. Not surprisingly, the strongest effects were observed for hypertension and essential hypertension, but there were also associations with ischemic heart disease and coronary atherosclerosis, myocardial infarction and angina pectoris, atrial fibrillation and flutter, subarachnoid hemorrhage and cerebral aneurysm, hypertensive heart disease and non-rheumatic valvular heart disease, as well as cardiac arrhythmias and paroxysmal tachycardia. Although classified under respiratory endpoints, there was also a significant association with lower odds of pulmonary edema. Additional outcomes with reduced odds included hypercholesterolemia and disorders of lipoprotein metabolism, sleep apnea and sleep-wake cycle disorders, disorders of the lacrimal system and corneal ulcer, salivary gland diseases, sensorineural hearing loss and ear disorders, as well as musculoskeletal outcomes, including limb pain, patella dislocation, vertebral fatigue fracture, and dorsalgia. Finally, there were isolated associations with psoriasis, kidney cyst, teeth disorders, a non-toxic single thyroid nodule, and iron deficiency anemia. In contrast, genetically proxied AGT downregulation was associated with higher odds of seven outcomes, which did not reveal any patterns. These outcomes included atrioventricular block, urinary retention, pneumonia, hernia, femur fracture, bladder cancer, and other white blood cell disorders.

**Figure 7.**
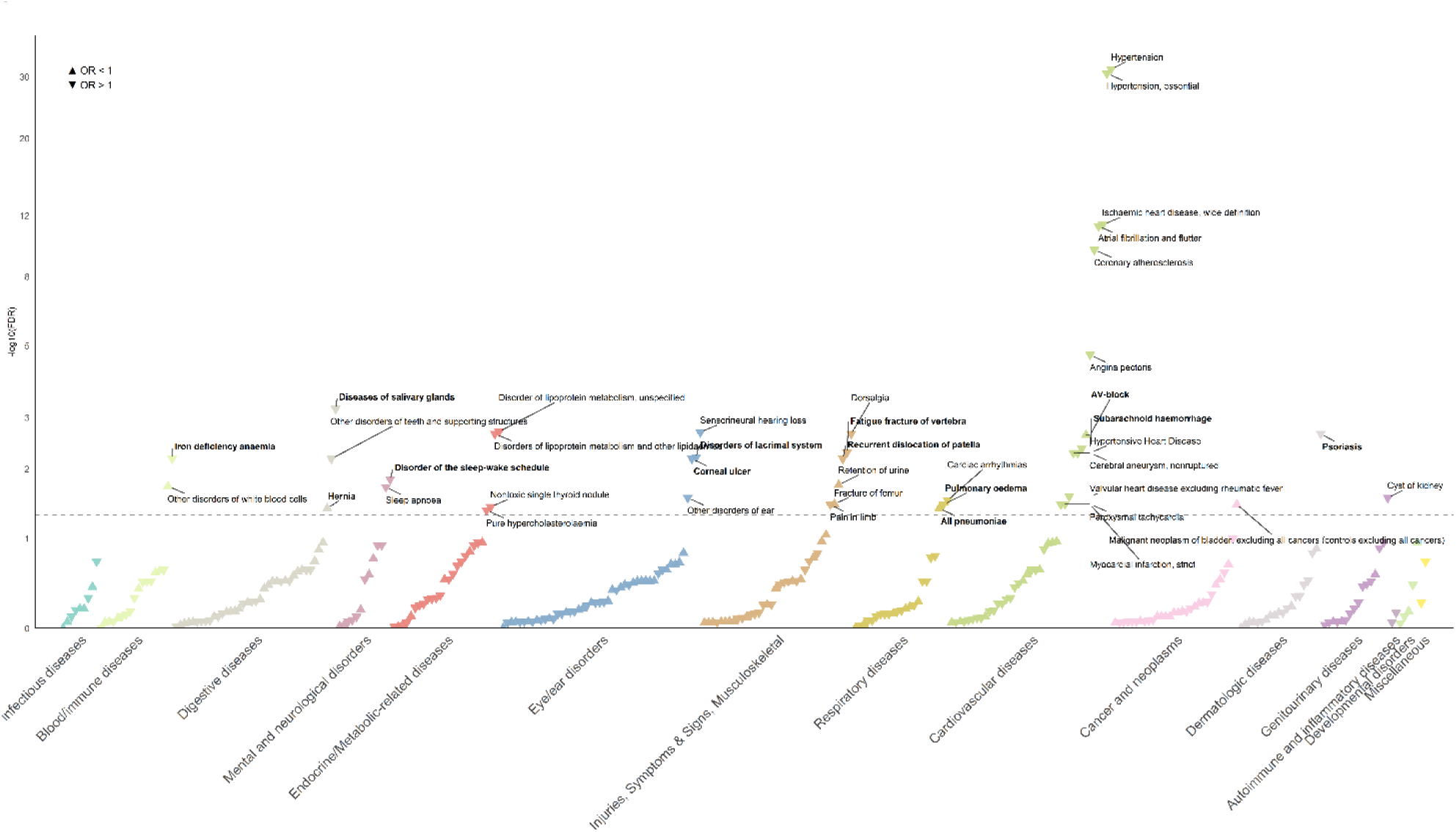
PheWAS results for genetically proxied angiotensinogen inhibition. The plot represents IVW MR association estimates with 326 binary clinical phenotypes. The downward and upward arrows represent associations with lower and higher outcome risk, respectively. All IVW - significant phenotypes after FDR correction (the dotted line represents FDR-adjusted p of 0.05) are annotated. The y axis corresponds to the negative 10-base logarithm of FDR-corrected p values, as derived from IVW MR analyses. OR – odds ratio, FDR – false discovery rate

## Discussion

Our drug-target Mendelian Randomization study supports that genetic variants mimicking the effect of pharmacological AGT synthesis inhibition are associated with a lower lifetime burden of cardiovascular disease. In line with experimental studies and clinical trial results, our genetic instruments were associated with lower systolic and diastolic blood pressure, as well as increases in blood potassium and renin levels. Genetically proxied SBP reduction via AGT downregulation was associated with lower risk of coronary artery disease, stroke and heart failure. These findings remained consistent in sensitivity analyses and when using complementary protein- and hepatic gene expression-lowering instruments. Furthermore, genetic evidence supports the non-inferiority of AGT synthesis inhibitors in lowering the risk of the three outcomes compared with standard-of-care RAAS-blocking antihypertensives.

Our results provide genetic support for the potential benefits of AGT synthesis inhibition across the spectrum of vascular disease. Genetically proxied SBP reduction due to perturbation in *AGT* was associated with lower risk of major CAD manifestations, including angina pectoris and MI, as well as lower CAC burden. The instrument was additionally associated with lower risk of ischemic stroke, including cardioembolic and small-vessel stroke, and subarachnoid hemorrhage, together with lower burden of imaging markers of cerebrovascular disease, such as WMH volume, mean diffusivity, and carotid plaque. Notably, we observed strong associations with MRI markers of cerebral small vessel disease and with lacunar stroke risk, supporting AGT inhibition as a promising strategy for slowing cerebral small vessel disease progression, a condition for which we currently lack disease-modifying therapies. In the HF domain, the instrument was associated with lower risk of all-cause HF and with favorable cardiac MRI endophenotypes, including lower LVEDV, LVM and LVmeanWT, which are markers of adverse ventricular remodeling and hypertrophy linked to HF risk^74,75^.

With respect to the safety-related outcomes, we observed a consistent signal for increases in blood potassium levels across all tested instruments and sensitivity analyses. In the PheWAS analysis, genetically proxied *AGT* synthesis inhibition was also associated with higher hyperkalemia risk, although the association did not persist after FDR correction. This finding is consistent with the expected physiological effects of RAAS inhibition and with observations from early-stage clinical trials of AGT synthesis inhibitors^18,19^. Whether hyperkalemia represents a clinically significant side-effect remains to be determined in phase 3 trials. However, phase 2 studies have reported only mild and transient hyperkalemia events that did not require treatment discontinuation. We also observed an isolated association between genetically proxied AGT level lowering and higher AST levels. This signal was not replicated with alternative instruments and was not accompanied by changes in ALT, an enzyme more specific for hepatocellular injury than AST^76^, or in other hepatic markers. It should be noted that our study design can only detect downstream effects of AGT synthesis perturbation rather than effects related to liver-targeting delivery strategies, such as GalNAc-based approaches. Importantly, we found no strong evidence for adverse effects on kidney function, as assessed by eGFR and UACR, and even observed a protective association for the binary endpoint of chronic kidney disease with our primary instrument. In sensitivity analyses using AGT proxies derived from SomaScan measurements, however, we observed reductions in both eGFR and UACR. These discordant findings warrant cautious interpretation and further evaluation in large-scale clinical trials.

Our phenome-wide analyses demonstrated associations with multiple hypertension- and atherosclerosis-related cardiovascular endpoints, thereby strengthening the findings from our drug-target MR analyses. Beyond cardiovascular disease outcomes, *AGT* downregulation was associated with lower risk of sensorineural hearing loss, a finding that may relate to hypertension^77^, which is increasingly recognized as a risk factor for age-related hearing loss, potentially through microangiopathy of the inner ear vasculature^78^. Additional associations were observed with disorders of the salivary and lacrimal glands as well as potentially related conditions such as corneal ulcer and dental disorders. These signals may reflect local RAAS activity, as expression of RAAS components, including *Agt*, has been detected in rodent lacrimal^79^ and salivary glands^80^. We also observed associations with established risk factors for cardiovascular disease, including sleep apnea and dyslipidemia. Sleep apnea is an established cause of secondary hypertension^81^, characterized by increased RAAS activity^82^ and is therefore more frequently screened for in patients with hypertension. Likewise, dyslipidemia is more commonly diagnosed in patients with hypertension and cardiovascular disease^83^. These signals may therefore reflect increased detection of cardiometabolic risk factors among individuals with hypertension rather than direct causal effects of *AGT* perturbation. With respect to safety signals, we reassuringly observed only a small number of isolated associations across the disease spectrum without clear clustering, suggesting that these findings may reflect rare or context-specific effects that warrant further investigation.

RNA-based therapeutics are increasingly being developed to treat cardiovascular disease, with their prolonged duration of action representing a key advantage. Besides AGT, these approaches aim at established cardiovascular targets, initiated or strongly motivated by human genetic evidence^84^, such as lipoprotein(a), Proprotein Convertase Subtilisin/Kexin type 9 (PCSK9), apolipoprotein C-III, angiopoietin-like 3 and coagulation factor XI^85,86^. The concurrent administration of multiple RNA-based therapeutics at intervals of several months, potentially in combination with the regular visit at the physician’s office, may improve the cardiometabolic profile and enhance adherence, thereby representing a promising strategy for primary and secondary cardiovascular event prevention.

Our study has specific limitations. First, although we used the largest available liver eQTL dataset (a meta-analysis of four studies), the relatively small sample size limits statistical power to detect additional hepatic gene expression-modifying variants at the *AGT* locus. Second, MR assesses the lifelong consequences of AGT synthesis inhibition, which may not reflect the short-term effects of pharmacological interventions. Third, despite using minimally correlated SNPs within the *AGT* locus, we cannot completely rule out potential effects on neighboring genes, horizontal pleiotropy or linkage disequilibrium. Fourth, although all variants in the main instrument were strongly associated with circulating AGT levels, the effects of most of these variants on blood pressure were modest, and the effects of the SBP- and DBP-weighted instruments in the UK Biobank were small. This selection could introduce weak instrument bias, which however would shift the effects towards the null^87^. Fifth, the majority of the population in the included GWAS was from European ancestry, potentially restricting the generalizability of our results to other ancestry groups.

In conclusion, genetic variation in *AGT* that mimics pharmacological inhibition of hepatic AGT synthesis, is associated with lower risk of cardiovascular disease and reduced burden of related etiologies, including atherosclerosis, cerebrovascular pathologies, and adverse cardiac remodeling. Our results provide human genetic evidence supporting the potential of angiotensinogen synthesis inhibition to meaningfully reduce cardiovascular risk and offer guidance for further clinical development.

## Acknowledgements

We acknowledge the Mega Vascular Cognitive Impairment and Dementia (MEGAVCID) consortium for providing us access to the vascular dementia GWAS summary statistics. Data for the UK Biobank analyses were accessed under application number 151281.

## Data availability statement

The GWAS Catalog study accession numbers or website links for the summary statistics used in our analyses are provided in **Supplementary Table 2.** The summary statistics for vascular dementia are available upon request to the MEGAVCID consortium, and UK Biobank data can be accessed upon submission of an application to the UK Biobank. All results generated in this study are available in **Supplementary Tables.**

## Appendices

Supplementary Figures 1-8

Supplementary Tables ST1-ST12

