## Supplementary Figures for "Genetically proxied inhibition of angiotensinogen synthesis is associated with lower cardiovascular risk"

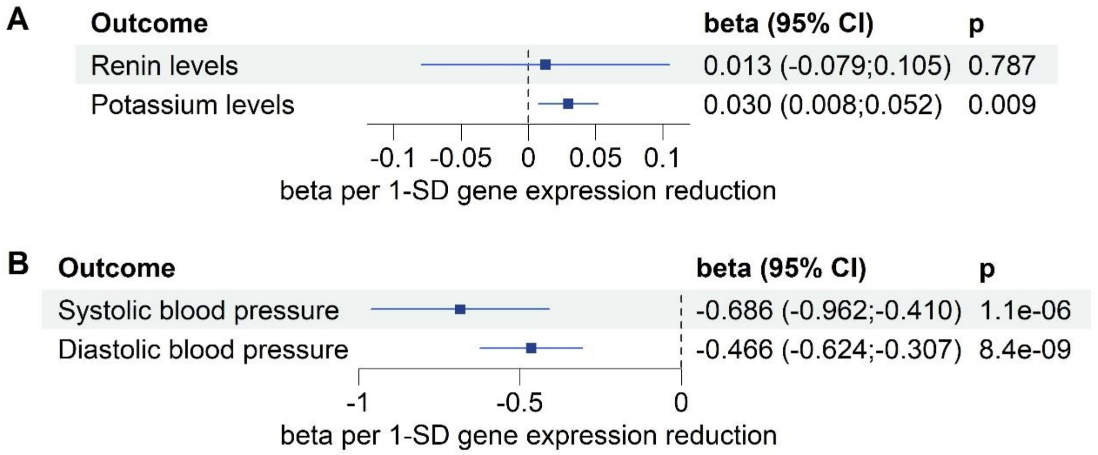


**Supplementary Figure 1. Positive control analysis for the gene expression instrument.** Forest plots depicting the IVW MR effects of the instrument on **(A)** circulating renin and potassium levels, **(B)** systolic and diastolic blood pressure. CI: confidence intervals, SD: standard deviation.

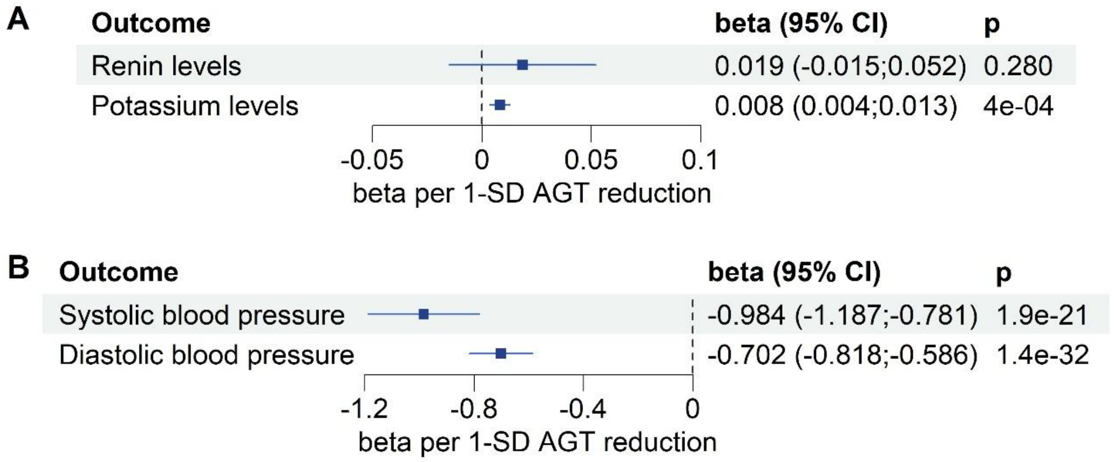


**Supplementary Figure 2. Positive control analysis for the SomaScan-based protein level instrument.** Forest plots depicting the IVW MR effects of the instrument on **(A)** circulating renin and potassium levels, **(B)** systolic and diastolic blood pressure. CI: confidence intervals, SD: standard deviation, AGT: angiotensinogen.

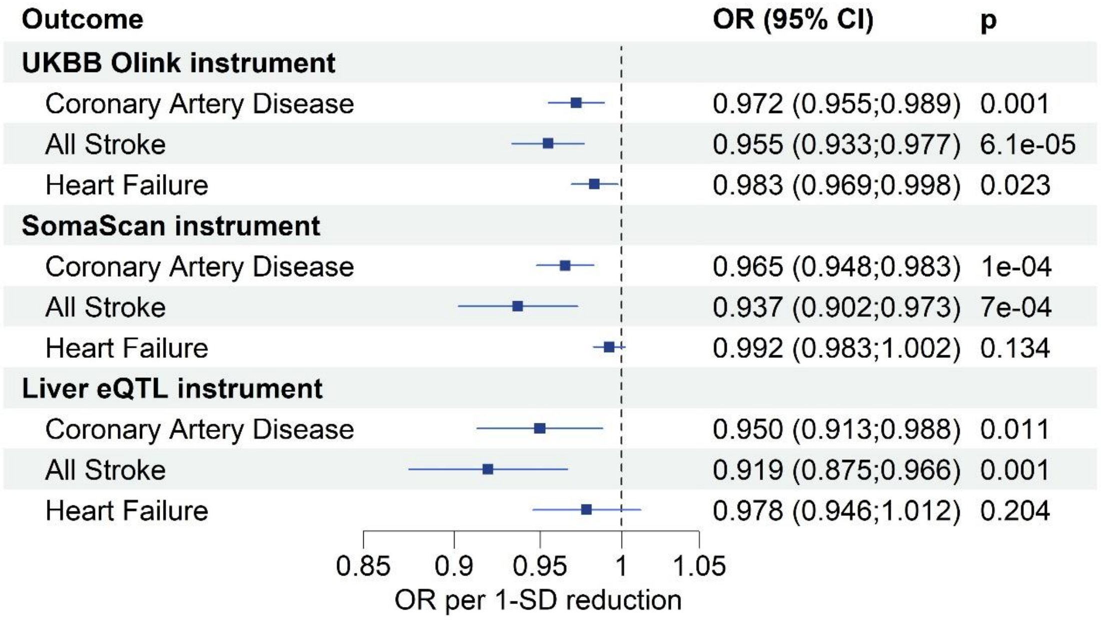


**Supplementary Figure 3. Mendelian Randomization results for primary outcomes.** Forest plot showing the IVW MR effects of the UKBB AGT instrument, the SomaScan AGT instrument and the gene expression instrument on CAD, stroke and heart failure risk. The derived odds ratios correspond to 1-SD reduction in circulating angiotensinogen levels or hepatic gene expression, respectively. OR – odds ratio, CI – confidence intervals, SD – standard deviation, UKBB – UK Biobank, eQTL – expressive quantitative trait loci

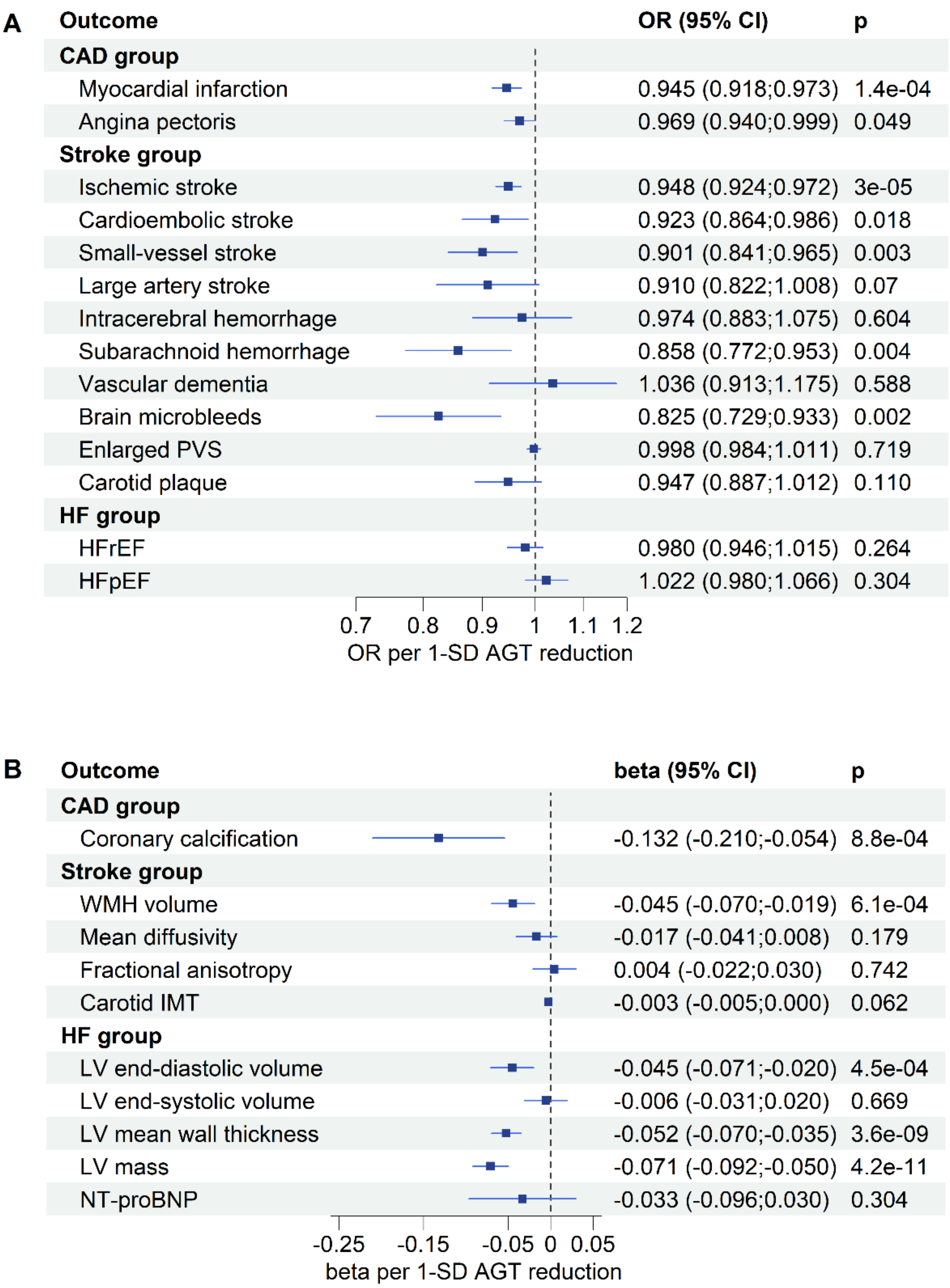


**Supplementary Figure 4. Mendelian Randomization results for secondary outcomes.** Forest plots showing the IVW MR effects of the UKBB angiotensinogen instrument on binary **(A)** and continuous **(B)** outcomes of the CAD, stroke and HF subgroups. The beta coefficients of the HF group continuous outcomes were scaled to represent 1-SD changes in each measurement. OR – odds ratio, CI – confidence intervals, SBP – systolic blood pressure, CAD – coronary artery disease, PVS – perivascular spaces, HF – heart failure, HFrEF – heart failure with reduced ejection fraction, HFpEF – heart failure with preserved ejection fraction, WMH – white matter hyperintensities, IMT – intima-media thickness, LV – left ventricle, NT-proBNP – N-terminal pro-B natriuretic peptide.

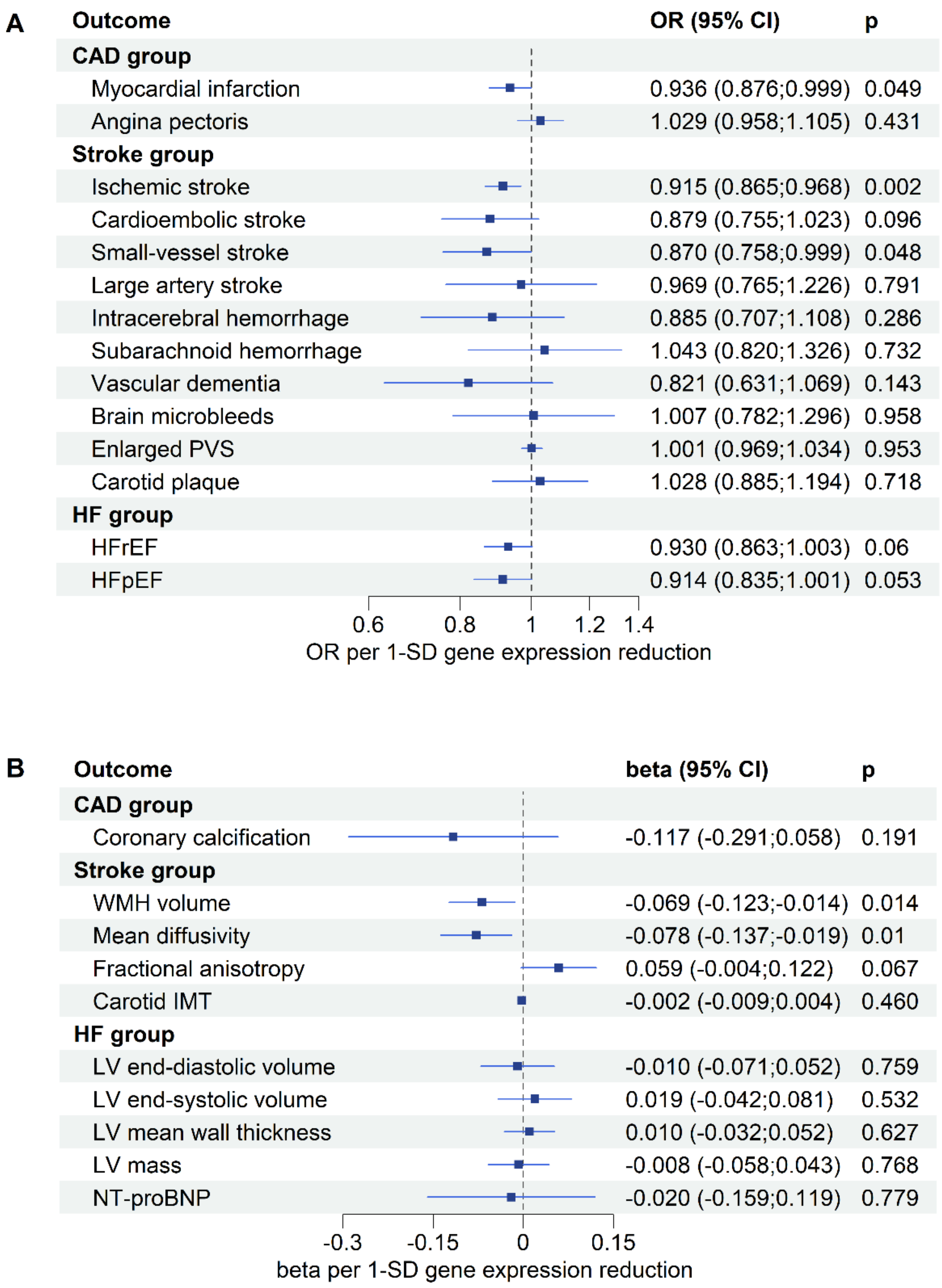


**Supplementary Figure 5. Mendelian Randomization results for secondary outcomes.** Forest plots showing the IVW MR effects of the gene expression instrument on binary **(A)** and continuous **(B)** outcomes of the CAD, stroke and HF subgroups. The beta coefficients of the HF group continuous outcomes were scaled to represent 1-SD changes in each measurement. OR – odds ratio, CI – confidence intervals, SBP – systolic blood pressure, CAD – coronary artery disease, PVS – perivascular spaces, HF – heart failure, HFrEF – heart failure with reduced ejection fraction, HFpEF – heart failure with preserved ejection fraction, WMH – white matter hyperintensities, IMT – intima-media thickness, LV – left ventricle, NT-proBNP – N-terminal pro-B natriuretic peptide.

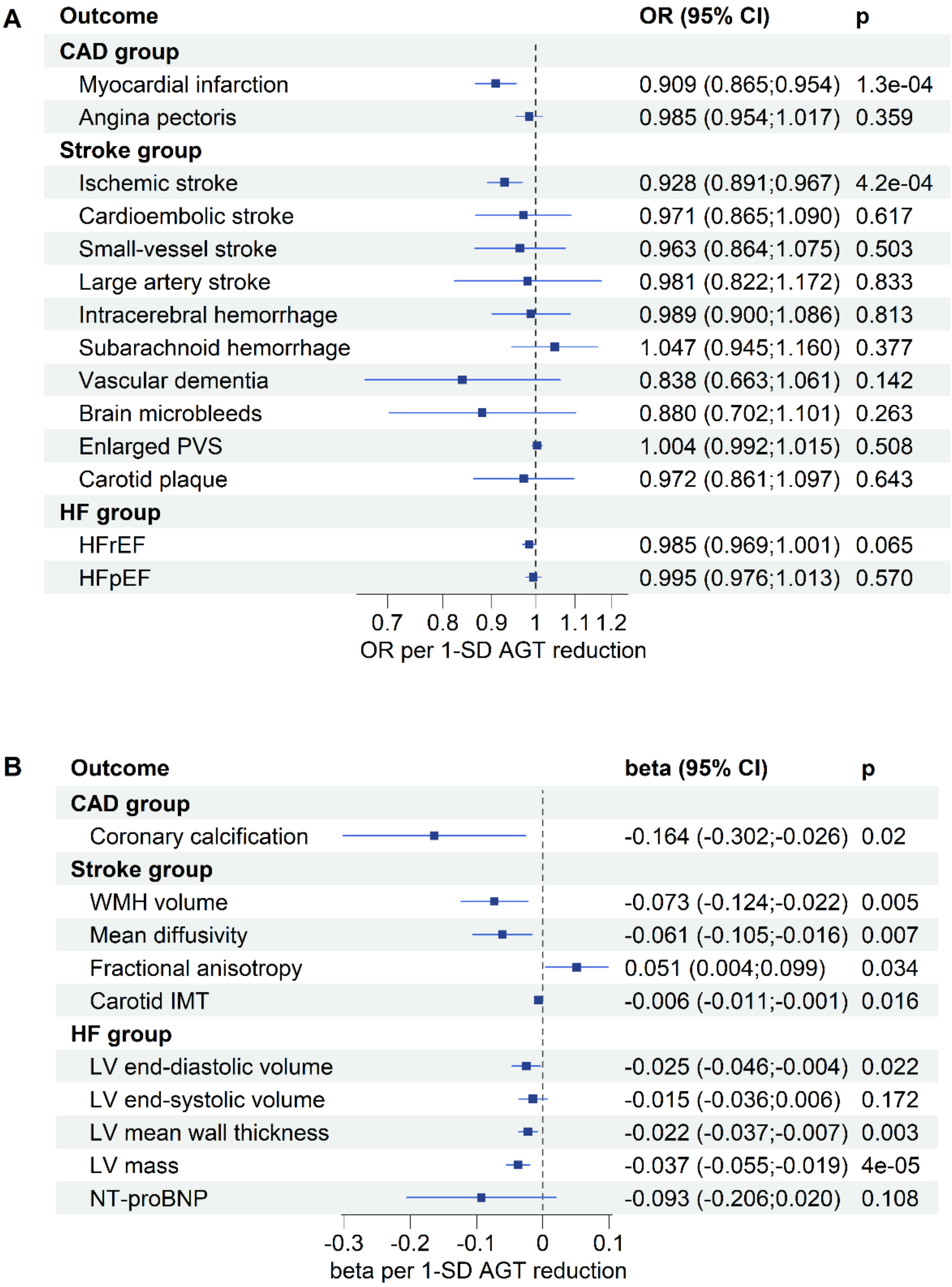


**Supplementary Figure 6. Mendelian Randomization results for secondary outcomes.** Forest plots showing the IVW MR effects of the SomaScan angiotensinogen instrument on binary **(A)** and continuous **(B)** outcomes of the CAD, stroke and HF subgroups. The beta coefficients of the HF group continuous outcomes were scaled to represent 1-SD changes in each measurement. OR – odds ratio, CI – confidence intervals, SBP – systolic blood pressure, CAD – coronary artery disease, PVS – perivascular spaces, HF – heart failure, HFrEF – heart failure with reduced ejection fraction, HFpEF – heart failure with preserved ejection fraction, WMH – white matter hyperintensities, IMT – intima-media thickness, LV – left ventricle, NT-proBNP – N-terminal pro-B natriuretic peptide.

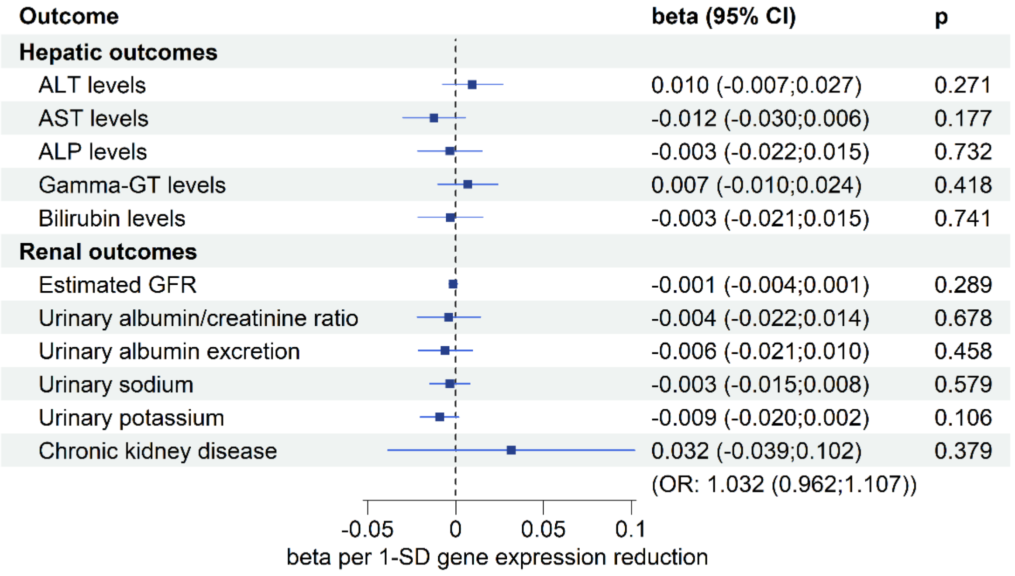


**Supplementary Figure 7. Mendelian Randomization results for safety-related outcomes.** Forest plot showing the IVW MR effects of the gene expression instrument on liver- and kidney-related parameters. OR – odds ratio, CI – confidence intervals, GFR – glomerular filtration rate, SD – standard deviation, AGT – angiotensinogen, ALT – alanine aminotransferase, AST – aspartate aminotransferase, ALP – alkaline phosphatase, Gamma-GT - gamma-glutamyl transferase.

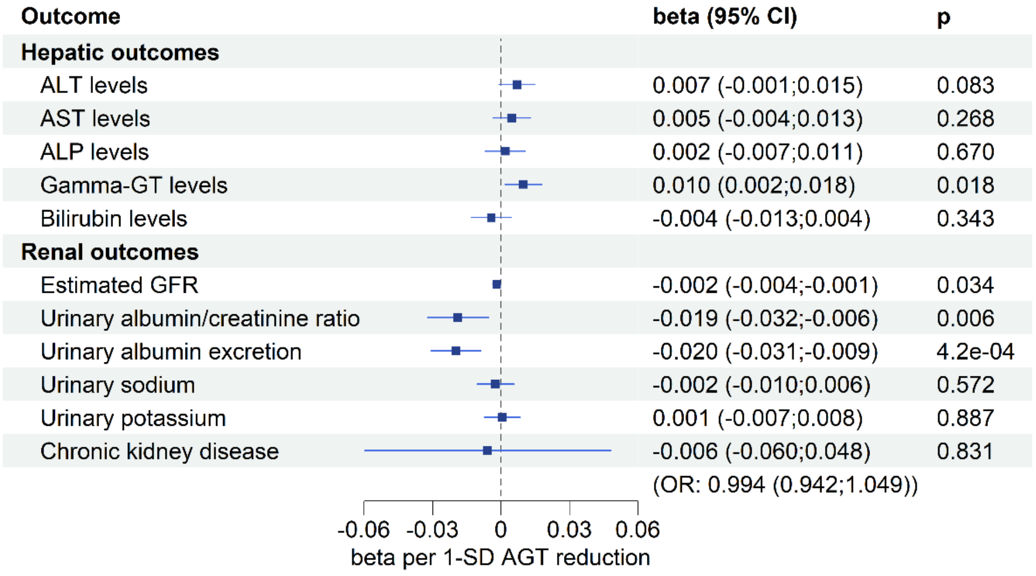


**Supplementary Figure 8. Mendelian Randomization results for safety-related outcomes.** Forest plot showing the IVW MR effects of the SomaScan AGT instrument on liver- and kidney-related parameters. OR – odds ratio, CI – confidence intervals, GFR – glomerular filtration rate, SD – standard deviation, AGT – angiotensinogen, ALT – alanine aminotransferase, AST – aspartate aminotransferase, ALP – alkaline phosphatase, Gamma-GT - gamma-glutamyl transferase.
